# Racial Differences and Contributory Cardiovascular and Non-cardiovascular Risk Factors Towards Chronic Kidney Disease Progression In Young To Middle-Aged Black And White American Adults

**DOI:** 10.1101/2023.02.13.23285888

**Authors:** Yuni Choi, David R Jacobs, Holly J. Kramer, Gautam R. Shroff, Alexander R. Chang, Daniel A Duprez

## Abstract

**Background:** The progression of chronic kidney disease (CKD) is higher in Black than in White Americans but studies have mainly focused on racial differences within advanced CKD. We evaluated CKD progression in Black and White participants over 20 years and the contribution of conventional cardiovascular and non-traditional risk factors to racial disparities in CKD progression.

**Methods:** This study was based on 2,175 Black and 2,207 White adults in the Coronary Artery Risk Development in Young Adults (CARDIA) study. Both estimated glomerular filtration rate (eGFR) and urinary albumin-to-creatinine ratio (UACR) were measured at study year 10 (age 27-41y) and every five years for 20 years. The outcome was CKD progression through No CKD into Low, Moderate, High, or Very High Risk that was based on categories of eGFR and UACR in combination. The association between race and CKD progression as well as the contribution of risk factors to racial differences were assessed in multivariable-adjusted Cox proportional hazards models.

**Results:** Black participants had higher CKD transition probabilities than White participants and more prevalent risk factors during the 20-year period studied. Hazard ratios for CKD transition for Black (vs White participants) were 1.38 from No CKD into ≥ Low Risk, 2.25 from ≤ Low Risk into ≥ Moderate Risk, and 4.49 for from ≤ Moderate Risk into ≥ High Risk. Racial differences in CKD progression from No CKD into ≥ Low Risk were primarily explained by forced vital capacity (54.8%), hypertension (30.9%), and obesity (20.8%). Similar findings were observed for the race difference in transition from ≤ Low Risk into ≥ Moderate Risk, but little of the race difference in transition ≤ Moderate Risk into ≥ High Risk was explained.

**Conclusions:** In this longitudinal study, Black compared to White participants had a higher risk of CKD progression, and this discrepancy may be partly explained by conventional cardiovascular and non-traditional risk factors.

**Clinical Perspective:** *What Is New?:*

▪ In 20 years of follow-up, young Black American adults had higher risk of chronic kidney disease (CKD) progression than their White counterparts, and the differences were larger in transitions to more advanced categories.
▪ Additionally, Black individuals had more conventional cardiovascular and non-traditional characteristics known to increase risk of CKD.

*What Are the Clinical Implications?:*

▪ Periodic screening for elevated albuminuria and eGFR would be helpful, particularly among young Black individuals.
▪ Future studies should evaluate if regular monitoring of eGFR and albuminuria in young Black individuals is helpful in preventing progression in CKD.

## Introduction

There are racial disparities in the prevalence of chronic kidney disease (CKD) in the US, with Black individuals suffering the highest frequency.^1^ Diabetes and hypertension are widely recognized risk factors for CKD and end stage kidney disease (ESKD).^2,3^ These disorders are the leading contributors to kidney failure in the US and together account for 64% of kidney failure.^4^ Another clinical risk factor for the onset of CKD that has been heavily researched is obesity.^5^ Although the exact causes of racial differences in CKD are not all understood, this likely in part reflects differences in the disproportionate prevalence of these major contributors.^6,7^ Social and economic disadvantages related to race or ethnicity may also generally help explain disparities in clinical conditions and CKD prevalence.^8^ There is currently little evidence from longitudinal data on the CKD progression rate between Black and White Americans from early adulthood to middle age, as determined by combinations of estimated glomerular filtration rate (eGFR) and urinary albumin-to-creatinine ratio (UACR). Furthermore, if racial disparities in CKD progression exist during this transitional period, little is known about the factors that contribute to such differences. Racial and ethnic differences exist in the rate of eGFR decline prior to the development of CKD, and traditional risk factors do not entirely explain these variations, calling for more research on this topic.^9^ We recently showed, using long term follow-up data from the Coronary Artery Risk Development in Young Adults (CARDIA) study that even minor deterioration in kidney function was associated with higher risk of incident CVD, total mortality, and the CKD progression through modified KDIGO categories;^10^ the CKD progression to worse CKD risk categories were predicted by conventional and non-traditional risk factors.^11^ Besides hypertension, diabetes, and obesity, which are well-known risk factors for CKD progression, we found that absolute lung function (as assessed by forced vital capacity (FVC)), markers of low-grade inflammation, serum urate, and the sum of 4 serum carotenoids also predicted CKD progression. All of these factors play a role in or are statistically associated with vascular disease and may therefore affect kidney function. Cross-talk between lung and kidney is well document.^12–14^ Lung and kidney endothelial tissue function are sensitive to microvascular changes.^15^ The kidney medulla has the lowest oxygen content in the body and is sensitive to minor hypoxemia, possibly leading to nephron loss and damage.^13^ Inflammation is a common thread in endothelial cell health.^16^ Urate may be a cause or a biomarker of CKD progression.^17–19^ The sum of 4 serum carotenoids (alpha- and beta-carotene, beta-cryptoxanthin, and zeaxanthin/ lutein) are empirically associated with diet quality, physical activity, smoking status, and body mass index^20^ and these carotenoids are strongly suggested to influence endothelial function.^21^

Building upon this evidence, herein, we prospectively examined CKD progression probabilities in Black and White as well as the contribution of conventional cardiovascular and non-traditional factors to racial differences in CKD progression. It was hypothesized that CKD progression rate would be higher among Black vs. White Americans and that the higher progression would be explained by disproportionate distribution of many clinical conditions.

## Methods

### Data Availability Statement

The data used in this study are available from the CARDIA Coordinating Center (www.cardia.dopm.uab.edu) on reasonable request.

### Study Design and population

This study was conducted in the CARDIA cohort, a longitudinal community-based study of Black and White men and women from four cities able to walk on a treadmill when they were 18 to 30 years old at the time of enrolment in 1985-1986 (exam Year 0).^22^ Furthermore, balanced representation of age, race (Black or White), sex, and educational attainment (<high school or ≥high school) was also a goal of the recruiting scheme at each of the field centers. CARDIA was approved by the institutional review boards of each study center, and at each follow-up visit, participants provided written informed consent. The current analyses composed of 4,382 participants after we excluded participants who had a history of cardiovascular disease or died before baseline for this study at Year 10 (n=90), had no kidney measurements (n=722), or had no data on hypertension, diabetes, dyslipidemia, or obese status at Year 10 (n=9).

### Assessment of kidney markers and definition of CKD progression

Serum creatinine, urinary albumin, and urinary creatinine were assayed up to 5 times between Year 10 and Year 30; eGFR was calculated using serum creatinine-based CKD-EPI 2021 race-free equation.^23,24^ Urinary albumin and creatinine were assessed with single untimed urinary specimens collected throughout the same exams. Hospitalized or fatal kidney failure was ascertained via annual surveys until Year 32 (August 31, 2018). The 2012 KDIGO heat map depicting CKD progression risk categories was modified to account for the low number of adults with large declines in eGFR, typical of a generally healthy younger group in CARDIA.^25^ The five CKD categories analyzed for this study were No CKD, Low Risk, Moderate Risk, High Risk, and Very High Risk (Figure S1). Specifically, 1) low UACR was split into two categories: <10 (mg/g) and 10-29 (as UACR 10-29 is common in early middle age), allowing distinction between No CKD from Low Risk with early increases in UACR; 2) we categorized the few individuals with eGFR 45-59 (mL/min/1.73 m^2^) and UACR <30 from Moderate Risk to High Risk to prevent mixing them with the participants with eGFR 45-59 (mL/min/1.73 m^2^) and UACR 10– 29. Other than that, the aforementioned categories and their labels correspond to KDIGO heat map categories.^25^ The highest CKD category for each exam was used, carrying forward the most recent non-missing value, if necessary. Therefore, sample sizes at Year 10, Year 15, Year 20, Year 25, and Year 30 were 3,461, 4,032, 4,261, 4,376, and 4,382 participants, respectively. The primary outcomes used in the current study were three binary transitions: Transition 1) from No CKD into Low, Moderate, High, or Very High Risk (or No CKD into ≥ Low Risk); Transition 2) from No CKD or Low Risk into Moderate, High, or Very High Risk (or ≤ Low Risk into ≥ Moderate Risk); and Transition 3) from No CKD, Low, or Moderate Risk into High or Very High Risk (or ≤ Moderate Risk into ≥ High Risk). The first and second outcomes primarily represent a rise in UACR, both while retaining eGFR ≥60.The third outcome represents transitions involving decreased eGFR, either alone or in conjunction with increase in UACR, or severely increased UACR with eGFR ≥60. Transitioning into the Very High Risk was not evaluated separately due to very few numbers.

### Assessment of covariates

We obtained information about age, self-reported race, and sex at Year 0 and at each exam have updated years of education completed, smoking status and pack-year of smoking, and medication use (diabetes, hypertension, and dyslipidemia) through self-reported questionnaire and/or review of medication bottles. Time-updated covariates were created for the current analysis. Measurements of all other physical and laboratory covariates were done between Year 10 and Year 30 except for several variables as described later. Body mass index (BMI, kg/m^2^) was measured, and obesity was defined as BMI ≥30 kg/m^2^. Hypertension was defined as systolic blood pressure≥140 mm Hg, diastolic blood pressure≥90 mm Hg, or the use of antihypertensive medication. Lung volume was measured as forced vital capacity (FVC; Year 0, Year 2, Year 5, Year 10, Year 20, and Year 30).^26^ Participants were asked to fast for 12 hours before each blood draw. Blood was drawn from the antecubital vein and serum and plasma aliquots were stored at −70°C until testing. Description of blood specimen collection and assay methodologies were described previously.^27–29^ Laboratory variables included triglycerides (Year 10-Year 30), HDL-C (Year 10-Year 30), serum urate (Year 10, Year 15, Year 20),^30^ high-sensitivity C-reactive protein (hsCRP; Year 7, Year 15, Year 20, Year 25),^31^ the nuclear magnetic resonance marker GlycA (Year 7),^32,33^ and sum of 4 carotenoids (alpha-carotene, beta-carotene, beta-cryptoxanthin, and zeaxanthin/lutein; Year 7, Year 15, Year 20).^34,35^ Other binary covariates included high triglycerides (serum triglycerides ≥150 mg/dL), low HDL-C (<40 for men and <50 mg/dL for women), and diabetes (fasting glucose concentration ≥126 mg/dL; 2-hour post-challenge glucose concentration ≥200 mg/dL, measured at Year 10, Year 20, and Year 25); glycated hemoglobin (HbA1c) ≥6.5%, measured at Year 20 and Year 25); and/or use of glucose lowering medication.^36^

### Statistical Analyses

We applied a 3-stage analytic approach to explore how potential risk factors for CKD progression differed between Black and White participants. First, we estimated the occurrence rates of transitions by race and then stratified by individual risk factors, presenting them as the percentage of people making three CKD transitions within extreme quartiles of risk factor categories. Second, minimal models adjusting for age, sex, and education were fitted using Cox proportional hazards regression; these models estimated the hazards ratios (HRs) of CKD transitions for Black vs White participants. Third, we then examined models further adjusting for each risk factor. The percentage change in the estimated log HR (or β, where e^β^ equals the HR for incident CKD transition in Black compared with White participants) were quantified to determine the extent to which the aforementioned risk factors contributed to racial differences in risk for CKD transition; the formula used is (β_race adjusted for predictor_ – β_race unadjusted_ / β_race unadjusted_) × 100. In a final step, mutually adjusted models that included all risk factors simultaneously were used to quantify the relative explanatory importance of combined risk factors for the same outcome prediction. Covariates were allowed to change with time. For continuous covariates, data were averaged over the available years up to each examination, and for the outcome of binary disease incidence (yes or no), the first disease occurrence was cumulative over study years of follow-up. Analytical strategies using time-varying covariates estimate the risk of CKD transition for each examination year, and the overall HR represents an average HR for the whole follow-up time. This approach carries forward in whole or in part the previously observed values and has the following advantage:^25^ 1) allowing for within-person variations in risk factor levels over the follow-up period, 2) relaxing the assumption of proportionality in which relative hazard remains constant over time with different predictors; and 3) retaining participants throughout follow-up. SAS version 9.4 (SAS Institute Inc., Cary, NC) was used for analyses, and two-sided P values <0.05 was considered statistically significant.

## Results

### Study Population Characteristics

The study included 4382 participants, of whom 2175 (49.6%) self-reported Black and 2207 self-reported White race (50.4%). For each race, the corresponding mean (SD) ages were 34.4 (3.8) and 35.6 (3.4) years. The baseline characteristics of the participants are presented in Table 1. Black participants in comparison to White participants were younger (34.4±3.8 vs. 35.6±3.4 years), fewer years of education (14.7±2.4 vs. 16.3±2.6 years), and a higher proportion were female (57.2 vs. 53.1%). In general, conventional and non-traditional characteristics were less favorable for Black individuals. For example, compared with White participants, Black participants tended to have lower levels and greater change of FVC and serum carotenoids, while GlycA, hsCRP, urate, hypertension, diabetes, and obesity were higher and/or increased more during follow-up. However, Black participants had lower levels of triglycerides.

**Table 1.** Baseline mean values of non-traditional and conventional cardiovascular risk factors according to race.

|  | Black participants |  | White participants |  | P-value |
| --- | --- | --- | --- | --- | --- |
|  | N | Mean±SD | N | Mean±SD |  |
| **Age at Year 10** | 2,175 | 34.3±3.8 | 2,207 | 35.6±3.4 | 0.02 |
| **Sex (Female), n (%)** | 2175 | 1245 (57.2) | 2207 | 1171 (53.1) | 0.005 |
| **Maximal educational level** | 2,175 | 14.7±2.4 | 2,207 | 16.3±2.6 | <0.001 |
| **Non-traditional risk factors** |  |  |  |  |  |
| **FVC** |  |  |  |  |  |
| Year 10 | 1,786 | 3.8±0.9 | 1,918 | 4.7±1.0 | <0.001 |
| Year 30 | 1,470 | 3.1±0.8 | 1,639 | 3.9±0.9 | <0.001 |
| Year 30- Year 10 | 1,248 | -0.7±0.4 | 1,472 | -0.7±0.4 | 0.76 |
| **Plasma GlycA** |  |  |  |  |  |
| Year 7 | 1,727 | 406.1±69.5 | 1,952 | 383.4±64.8 | <0.001 |
| **ln(serum hsCRP)** |  |  |  |  |  |
| Year 7 | 1,742 | 1.1±0.8 | 1,964 | 0.9±0.7 | <0.001 |
| Year 25 | 1,621 | 1.3±0.8 | 1,845 | 0.9±0.6 | <0.001 |
| Year 25-Year 7 | 1,343 | 0.2±0.8 | 1,678 | 0.04±0.7 | <0.001 |
| **Serum urate** |  |  |  |  |  |
| Year 0 | 2,139 | 5.1±1.4 | 2,191 | 5.4±1.4 | <0.001 |
| Year 20 | 1,401 | 5.8±1.6 | 1,736 | 5.7±1.4 | 0.02 |
| Year 20-Year 0 | 1,389 | 0.8±1.3 | 1,726 | 0.3±1.1 | <0.001 |
| **Sum of 4 serum carotenoids** |  |  |  |  |  |
| Year 7 | 1,672 | 44.9±22.9 | 1,887 | 54.1±30.3 | <0.001 |
| Year 20 | 1,219 | 44.0±24.9 | 1,509 | 53.9±33.2 | <0.001 |
| Year 20-Year 7 | 992 | -1.1±22.8 | 1,338 | 0.5±28.5 | 0.14 |
| **Conventional cardiovascular risk factors** |  |  |  |  |  |
| **Pack-year of smoking ≥10, n (%)** |  |  |  |  |  |
| Year 10 | 2,175 | 280 (12.9) | 2,207 | 391 (17.7) | <0.001 |
| Year 30 | 2,175 | 519 (23.9) | 2,207 | 480 (21.8) | 0.10 |
| Year 30-Year 10 | 2,175 | 239 (11.0) | 2,207 | 89 (4.0) | <0.001 |
| **Hypertension (≥140/90 mm Hg/**<br>**antihypertensive medications), n (%)** |  |  |  |  |  |
| Year 10 | 2,175 | 364 (16.7) | 2,207 | 162 (7.3) | <0.001 |
| Year 30 | 2,175 | 1,214 (55.8) | 2,207 | 682 (30.9) | <0.001 |
| Year 30-Year 10 | 2,175 | 850 (39.1) | 2,207 | 520 (23.6) | <0.001 |
| **Diabetes (plasma glucose $\geq 126$ mg/dL, 2-hr glucose $\geq 200$ mg/dL, or Hb1Ac $\geq 6.5\%$/antidiabetic medications), n (%)** |  |  |  |  |  |
| Year 10 | 2,175 | 69 (3.2) | 2,207 | 44 (2.0) | 0.01 |
| Year 30 | 2,175 | 446 (20.5) | 2,207 | 231 (10.5) | <0.001 |
| Year 30-Year 10 | 2,175 | 377 (17.3) | 2,207 | 187 (8.5) | <0.001 |
| **High plasma triglycerides ($\geq 150$ mg/dL), n (%)** |  |  |  |  |  |
| Year 10 | 2,175 | 306 (14.1) | 2,207 | 537 (24.3) | <0.001 |
| Year 30 | 2,175 | 626 (28.8) | 2,207 | 943 (42.7) | <0.001 |
| Year 30-Year 10 | 2,175 | 320 (14.7) | 2,207 | 406 (18.4) | 0.001 |
| **Low HDL-C (plasma $< 50$ mg/dL for female and $< 40$ mg/dL for male), n (%)** |  |  |  |  |  |
| Year 10 | 2,175 | 1211 (55.7) | 2,207 | 1244 (56.4) | 0.65 |
| Year 30 | 2,175 | 1377 (63.3) | 2,207 | 1389 (62.9) | 0.80 |
| Year 30-Year 10 | 2,175 | 320 (14.7) | 2,207 | 406 (18.4) | 0.001 |
| **Obesity ($\geq 30$ kg/m<sup>2</sup>), n (%)** |  |  |  |  |  |
| Year 10 | 2,175 | 834 (38.3) | 2,207 | 427 (19.4) | <0.001 |
| Year 30 | 2,175 | 1338 (61.5) | 2,207 | 892 (40.4) | <0.001 |
| Year 30-Year 10 | 2,175 | 504 (23.2) | 2,207 | 465 (21.1) | 0.09 |

FVC, forced vital capacity; HDL-C, high-density lipoprotein cholesterol; hsCRP, high-sensitivity C-reactive protein; ln, natural logarithm; SD, standard deviation.

### CKD transition probability by race

Black individuals had greater probabilities for all three transitions compared to White participants across the 20-year period (Table 2). The transition probabilities for Black and White participants were 33% vs 25.5% for No CKD into ≥ Low Risk, 15.9% vs 8.1% for ≤ Low Risk into ≥ Moderate Risk, and 8.1% vs 2.0% for ≤ Moderate Risk into ≥ High Risk. The non-traditional factors FVC and sum of 4 carotenoids showed inverse associations with advancing into higher categories when comparing extreme quartiles, while GlycA, hsCRP, and serum urate had positive associations. The only exception to the observed direction of association was for FVC in Black participants in the transition from ≤ Moderate Risk into ≥ High Risk. Of conventional factors, transitioning into higher categories was most strongly associated with diabetes and hypertension in both Black and White participants, but associations with cigarette smoking were weak. In some instances, the patterns were nominally more pronounced among the White participants.

**Table 2.**
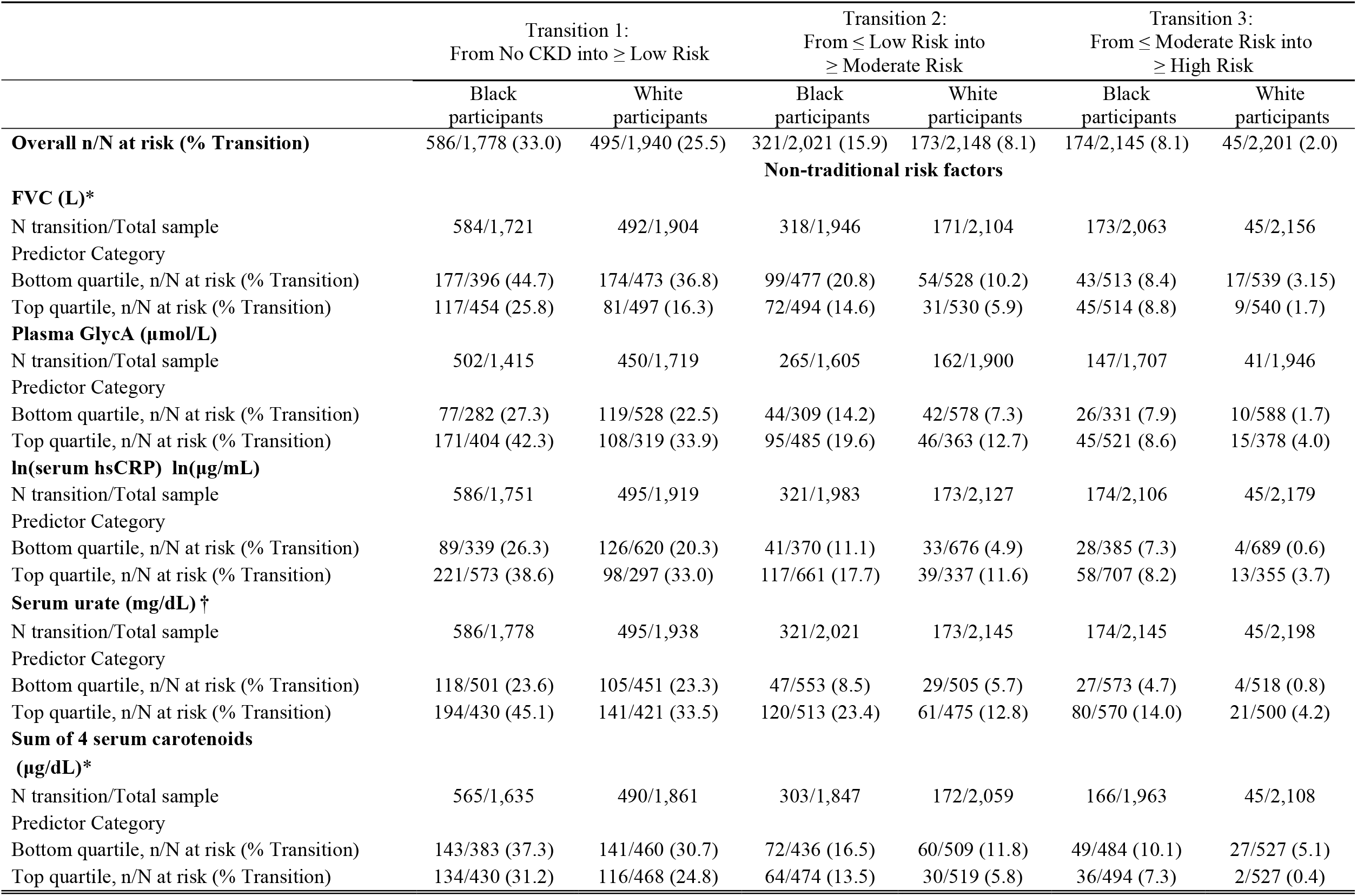
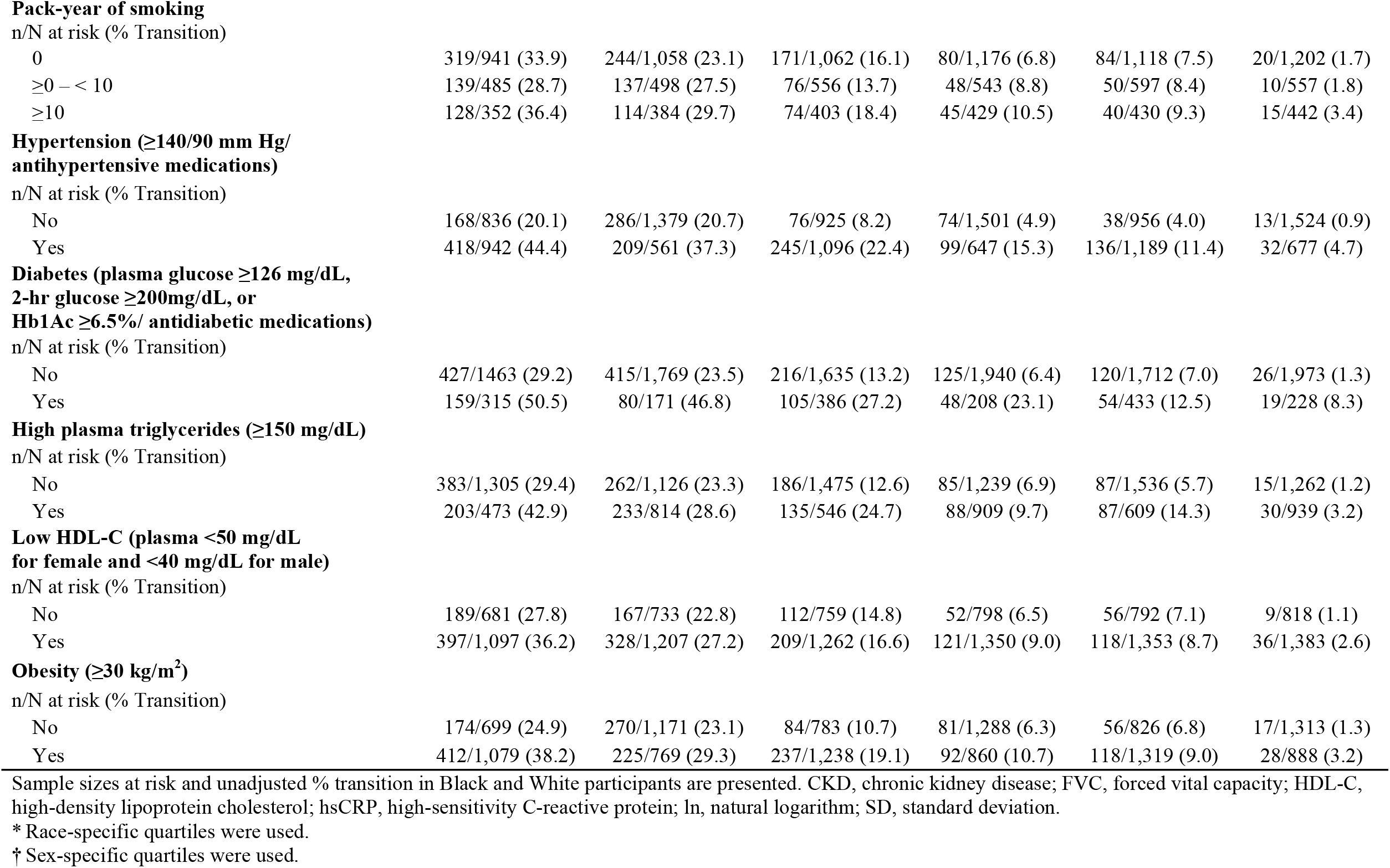
CKD transition probabilities according to non-traditional and conventional cardiovascular risk factors.

### Contribution of risk factors to the association between Black race and CKD progression

Table 3 shows the adjusted HRs for CKD progression for Black participants compared to White participants using the minimal model (age, sex, and education adjusted) and percent change in the β estimates after additional individual risk factor adjustment relative to the minimal model in proportional hazards regression. Black participants had higher risk of CKD progression, with the association being stronger with more severe transitions; the adjusted HR in Black participants compared with White participants were 1.38 for No CKD into ≥ Low Risk, 2.25 for ≤ Low Risk into ≥ Moderate Risk, and 4.49 for ≤ Moderate Risk into ≥ High Risk. In a series of analyses associated with each single risk factor, the dominant variables explaining of racial differences in CKD progression were FVC, followed by hypertension and obesity. Racial differences in FVC accounted for 54.8%, 32.2% and 14.2 % of the racial differences in the three transitions, respectively. The corresponding estimates were 30.9%, 20.1%, and 13.7% for hypertension, and 20.8%, 12.5%, and 4.7% for obesity. Some other factors explained smaller amounts of the race coefficient, and adjustment for TG enhanced the race coefficient. Simultaneous adjustment for all risk factors had a similar result as for adjustment only for FVC. In general, the transition to the highest categories (≤ Moderate Risk into ≥ High Risk) was less explained by risk factors studied compared with the first two transitions.

**Table 3.**
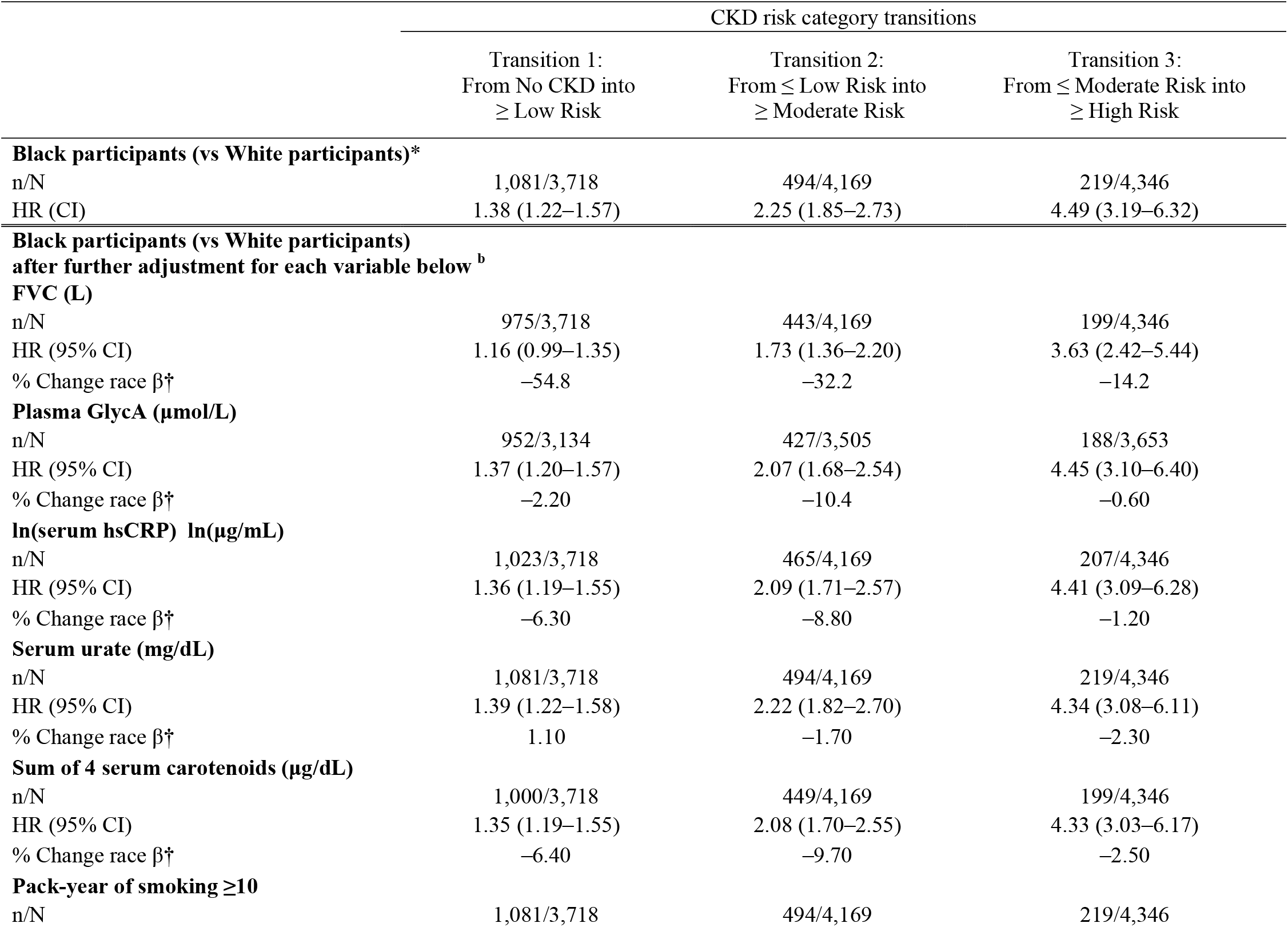
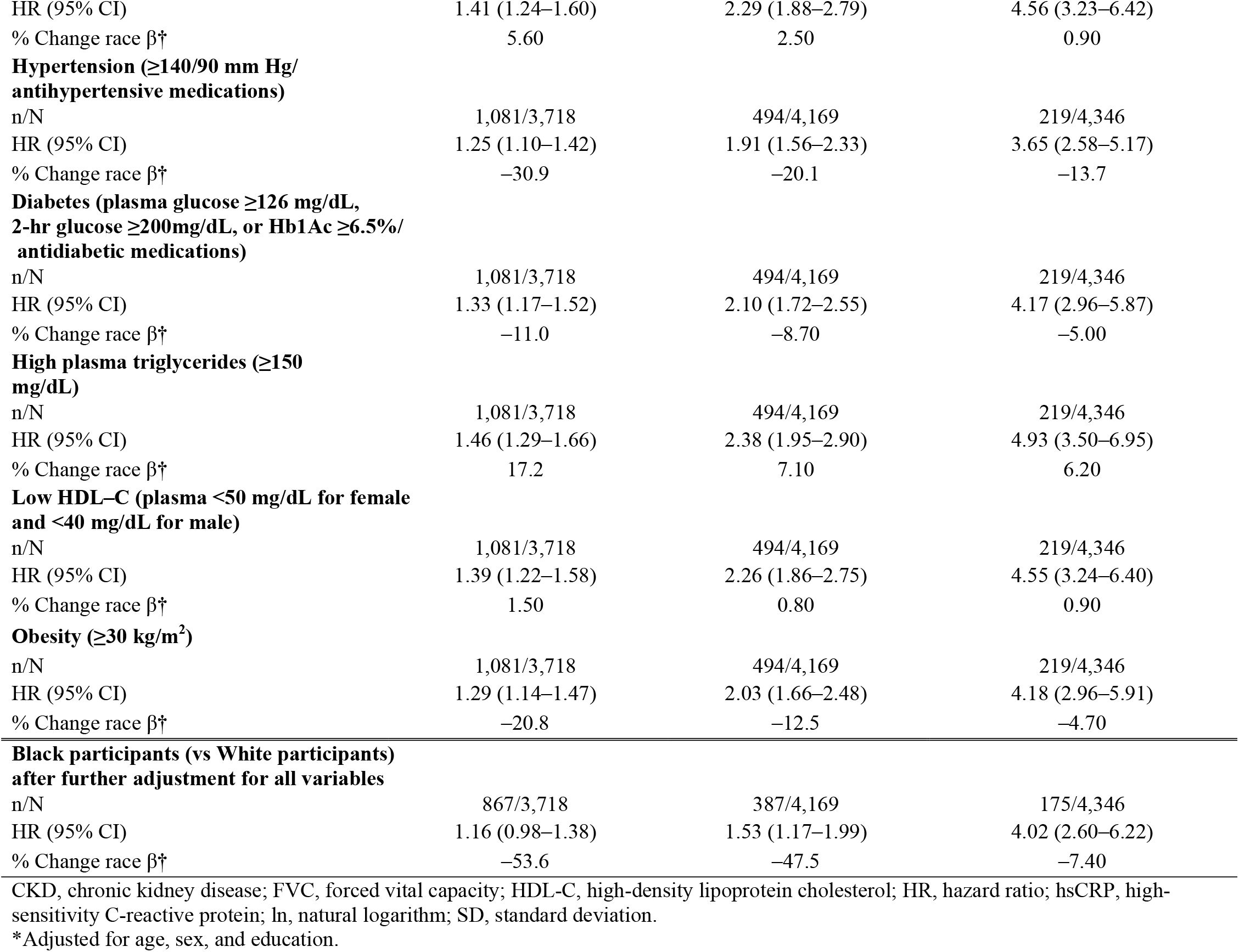
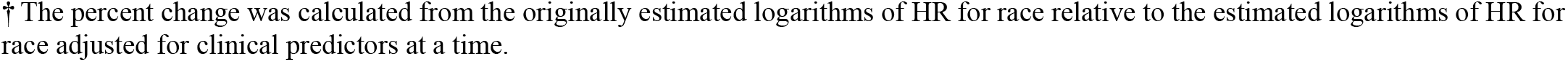
Adjusted HRs of CKD progression associated with Black participants (vs White participants), adjusted for time-varying predictors, one at a time and simultaneously.

|  | CKD risk category transitions |  |  |
| --- | --- | --- | --- |
|  | Transition 1:<br>From No CKD into<br>≥ Low Risk | Transition 2:<br>From ≤ Low Risk into<br>≥ Moderate Risk | Transition 3:<br>From ≤ Moderate Risk into<br>≥ High Risk |
| **Black participants (vs White participants)\*** |  |  |  |
| n/N | 1,081/3,718 | 494/4,169 | 219/4,346 |
| HR (CI) | 1.38 (1.22–1.57) | 2.25 (1.85–2.73) | 4.49 (3.19–6.32) |
| **Black participants (vs White participants)**<br>**after further adjustment for each variable below <sup>b</sup>** |  |  |  |
| **FVC (L)** |  |  |  |
| n/N | 975/3,718 | 443/4,169 | 199/4,346 |
| HR (95% CI) | 1.16 (0.99–1.35) | 1.73 (1.36–2.20) | 3.63 (2.42–5.44) |
| % Change race $\beta^\dagger$ | –54.8 | –32.2 | –14.2 |
| **Plasma GlycA (μmol/L)** |  |  |  |
| n/N | 952/3,134 | 427/3,505 | 188/3,653 |
| HR (95% CI) | 1.37 (1.20–1.57) | 2.07 (1.68–2.54) | 4.45 (3.10–6.40) |
| % Change race $\beta^\dagger$ | –2.20 | –10.4 | –0.60 |
| **ln(serum hsCRP) ln(μg/mL)** |  |  |  |
| n/N | 1,023/3,718 | 465/4,169 | 207/4,346 |
| HR (95% CI) | 1.36 (1.19–1.55) | 2.09 (1.71–2.57) | 4.41 (3.09–6.28) |
| % Change race $\beta^\dagger$ | –6.30 | –8.80 | –1.20 |
| **Serum urate (mg/dL)** |  |  |  |
| n/N | 1,081/3,718 | 494/4,169 | 219/4,346 |
| HR (95% CI) | 1.39 (1.22–1.58) | 2.22 (1.82–2.70) | 4.34 (3.08–6.11) |
| % Change race $\beta^\dagger$ | 1.10 | –1.70 | –2.30 |
| **Sum of 4 serum carotenoids (μg/dL)** |  |  |  |
| n/N | 1,000/3,718 | 449/4,169 | 199/4,346 |
| HR (95% CI) | 1.35 (1.19–1.55) | 2.08 (1.70–2.55) | 4.33 (3.03–6.17) |
| % Change race $\beta^\dagger$ | –6.40 | –9.70 | –2.50 |
| **Pack-year of smoking ≥10** |  |  |  |
| n/N | 1,081/3,718 | 494/4,169 | 219/4,346 |
| HR (95% CI) | 1.41 (1.24–1.60) | 2.29 (1.88–2.79) | 4.56 (3.23–6.42) |
| % Change race $\beta^\dagger$ | 5.60 | 2.50 | 0.90 |
| **Hypertension ($\geq 140/90$ mm Hg/**<br>**antihypertensive medications)** |  |  |  |
| n/N | 1,081/3,718 | 494/4,169 | 219/4,346 |
| HR (95% CI) | 1.25 (1.10–1.42) | 1.91 (1.56–2.33) | 3.65 (2.58–5.17) |
| % Change race $\beta^\dagger$ | –30.9 | –20.1 | –13.7 |
| **Diabetes (plasma glucose $\geq 126$ mg/dL,**<br>**2-hr glucose $\geq 200$ mg/dL, or Hb1Ac $\geq 6.5\%$/**<br>**antidiabetic medications)** |  |  |  |
| n/N | 1,081/3,718 | 494/4,169 | 219/4,346 |
| HR (95% CI) | 1.33 (1.17–1.52) | 2.10 (1.72–2.55) | 4.17 (2.96–5.87) |
| % Change race $\beta^\dagger$ | –11.0 | –8.70 | –5.00 |
| **High plasma triglycerides ($\geq 150$**<br>**mg/dL)** |  |  |  |
| n/N | 1,081/3,718 | 494/4,169 | 219/4,346 |
| HR (95% CI) | 1.46 (1.29–1.66) | 2.38 (1.95–2.90) | 4.93 (3.50–6.95) |
| % Change race $\beta^\dagger$ | 17.2 | 7.10 | 6.20 |
| **Low HDL-C (plasma $< 50$ mg/dL for female**<br>**and $< 40$ mg/dL for male)** |  |  |  |
| n/N | 1,081/3,718 | 494/4,169 | 219/4,346 |
| HR (95% CI) | 1.39 (1.22–1.58) | 2.26 (1.86–2.75) | 4.55 (3.24–6.40) |
| % Change race $\beta^\dagger$ | 1.50 | 0.80 | 0.90 |
| **Obesity ($\geq 30$ kg/m<sup>2</sup>)** |  |  |  |
| n/N | 1,081/3,718 | 494/4,169 | 219/4,346 |
| HR (95% CI) | 1.29 (1.14–1.47) | 2.03 (1.66–2.48) | 4.18 (2.96–5.91) |
| % Change race $\beta^\dagger$ | –20.8 | –12.5 | –4.70 |
| **Black participants (vs White participants)**<br>**after further adjustment for all variables** |  |  |  |
| n/N | 867/3,718 | 387/4,169 | 175/4,346 |
| HR (95% CI) | 1.16 (0.98–1.38) | 1.53 (1.17–1.99) | 4.02 (2.60–6.22) |
| % Change race $\beta^\dagger$ | –53.6 | –47.5 | –7.40 |

CKD, chronic kidney disease; FVC, forced vital capacity; HDL-C, high-density lipoprotein cholesterol; HR, hazard ratio; hsCRP, high-sensitivity C-reactive protein; ln, natural logarithm; SD, standard deviation.
\*Adjusted for age, sex, and education.
† The percent change was calculated from the originally estimated logarithms of HR for race relative to the estimated logarithms of HR for race adjusted for clinical predictors at a time.

## Discussion

The novelty of our study is that we demonstrated large differences between young Black and White Americans in risk of CKD progression across younger adulthood and quantified potential underlying markers and mechanisms linking race to CKD progression. In this long-term longitudinal study, Black individuals had a greater probability of progressing to more advanced CKD categories, and the observed race association became stronger in more severe transitions, for example in the 4.5-fold higher risk of CKD among Blacks vs. Whites for the transition from ≤ Moderate Risk into ≥ High Risk. We also found that Black participants had higher incidence of individual risk factors that predict CKD progression. Particularly, FVC, hypertension, and obesity were major factors that accounted for the racial differences in CKD transitions, with an earlier transition being better explained by these factors. The rest of the traditional and non-conventional factors across younger adulthood variably explained smaller magnitudes of the racial differences in CKD progression, and, in particular, FVC provided a weaker explanation than many of the other predictors of the race difference in the most severe transition.

This study complements previous findings^1,37,38^ by demonstrating racial differences in longitudinal CKD progression from young to middle adulthood using modified KDIGO categories that incorporated eGFR and UACR in combination. In our study, racial differences in CKD progression were mainly explained by absolute FVC, a non-conventional risk factor for CKD progression,^11^ with considerable differences in CKD progression probabilities for high vs low FVC. The reasons underlying racial differences in FVC remain unresolved but prior studies suggested that height and frame size may be more important to explain these differences than socioeconomic, nutritional, and environmental factors.^39,40^

Consistent with our findings, higher burden of chronic disease and earlier onset of multimorbidity were observed in Black than in White middle-aged Americans.^41^ Hypertension, diabetes, and obesity are intertwined (cardiometabolic syndrome) and are well-known risk factors for CKD and ESKD.^2,3,5^ These traditional risk factors at younger age were strongly associated with CKD progression in a previous CARDIA analysis.^11^ In a nationally representative sample, the prevalence of CKD decreased from 2003-2006 to 2015-2018 within the high-risk group for diabetes and CVD, but rising rates of obesity, hypertension, and diabetes offset all progress achieved through the improvement of CKD rate, underscoring the importance of addressing these important risk factors in an effort to lower the overall burden of CKD.^42^ Development of cardiometabolic dysfunction early in life among Black Americans may indicate earlier and longer cumulative exposure to risk factors that contribute to higher risk of CKD progression.^43,44^ Overall, a set of individual risk factors studied could account for about half of the racial difference in less severe CKD progression and, to a smaller extent, in more severe CKD transitions. In parallel to the clinical factors, a number of social components that are complexly interrelated (so called systemic racism) may also comprise a part of an explanation for racial differences in CKD progression: health services (e.g., access to and quality of care and insurance status), physical environment (e.g., housing quality and stability, neighborhood, and transportation), social environment (e.g., educational level, income level, and discrimination), social support, and health literacy.^8,45^ Our findings add to published findings in CARDIA delineating several ways in which social determinants of health and clinical factors contribute to Black-White differences in risky behaviors^46^, obesity,^47^ and CVD.^48^ Moreover, racial disparities in CKD may be explained by variants in the APOL1 gene that were known to link with end stage kidney disease and CKD in Black people, but rarely occurs in White people. In previous CARDIA work, in fully adjusted analysis, the estimated the population attributable fraction of albuminuria incidence explained by APOL1 high-risk genotype was 10.8% among Black participants.^49^ The remaining difference in CKD progression could be attributable to unmeasured factors or other biological, genetic, environmental factors. Further research is needed to better understand contributor to racial differences in CKD progression.

The strengths of our study were first, CARDIA is a community-based, long-term follow-up longitudinal study with a high follow-up rate among survivors. Second, a wide range of clinical variables were assessed and repeatedly measured many times, allowing for the calculation of the average effect over 20 years. Third, CKD was identified based on a modified KDIGO 2012 classification that suggested using eGFR and UACR in combination, and dynamic progression across CKD categories with non-traditional and conventional factors was evaluated. Lastly, our findings for FVC, which substantially explained the relationship between race and CKD progression, were novel and can provide further insight into a factor yielding potential racial differences in CKD progression. This study also has several limitations. The observational nature of this study precludes causal inference and residual confounding may exist. Additionally, CARDIA included only Black and White Americans, other races were not represented.

The findings of our longitudinal study suggest that CKD progression is more common in Black than White counterparts during the transition from young to middle-aged adulthood. Moreover, these differences may be explained partly by key non-traditional and conventional risk factors that showed apparent differences in CKD transition probabilities between them, notably FVC, hypertension, and obesity. Important factors in avoiding CKD progression include hypertension, diabetes, absolute FVC, and obesity.^11^ In this paper, we showed that FVC, hypertension, and obesity are dominant in explaining the race difference in CKD progression and both were more strongly related to UACR changes than to eGFR changes. Preserving kidney function is important for all persons, regardless of their race. However, given the disproportionate burden of risk factors for CKD among Black adults, it is particularly important to address the risk factors for CKD in Black adults in order to reduce disparities in CKD and its downstream consequences including end stage renal disease and cardiovascular disease. Because CKD progression can begin in younger adulthood and is attributed to various adverse health conditions, the implication is that primordial prevention should begin early in adult life. To close the equity gap related to the foremost risk factors for CKD and burden of CKD, strategic actions are needed to eliminate the determinants of population health known to increase risk of CKD progression and improve the health of individuals, families, and communities in the context of racial disparities.

Currently there are no recommendations for CKD screening by the U.S. Preventive Task Force. Blood pressure and BMI measurement are currently recommended for screening in young Black people every year. Assessment of eGFR and albuminuria should also be considered, especially if other known risk factors for CKD are present such as diabetes or hypertension.

## Data Availability

Data used in this paper are available on reasonable request from the CARDIA Coordinating Center, https://www.cardia.dopm.uab.edu/

## Supplemental Material

**Figure S1.**
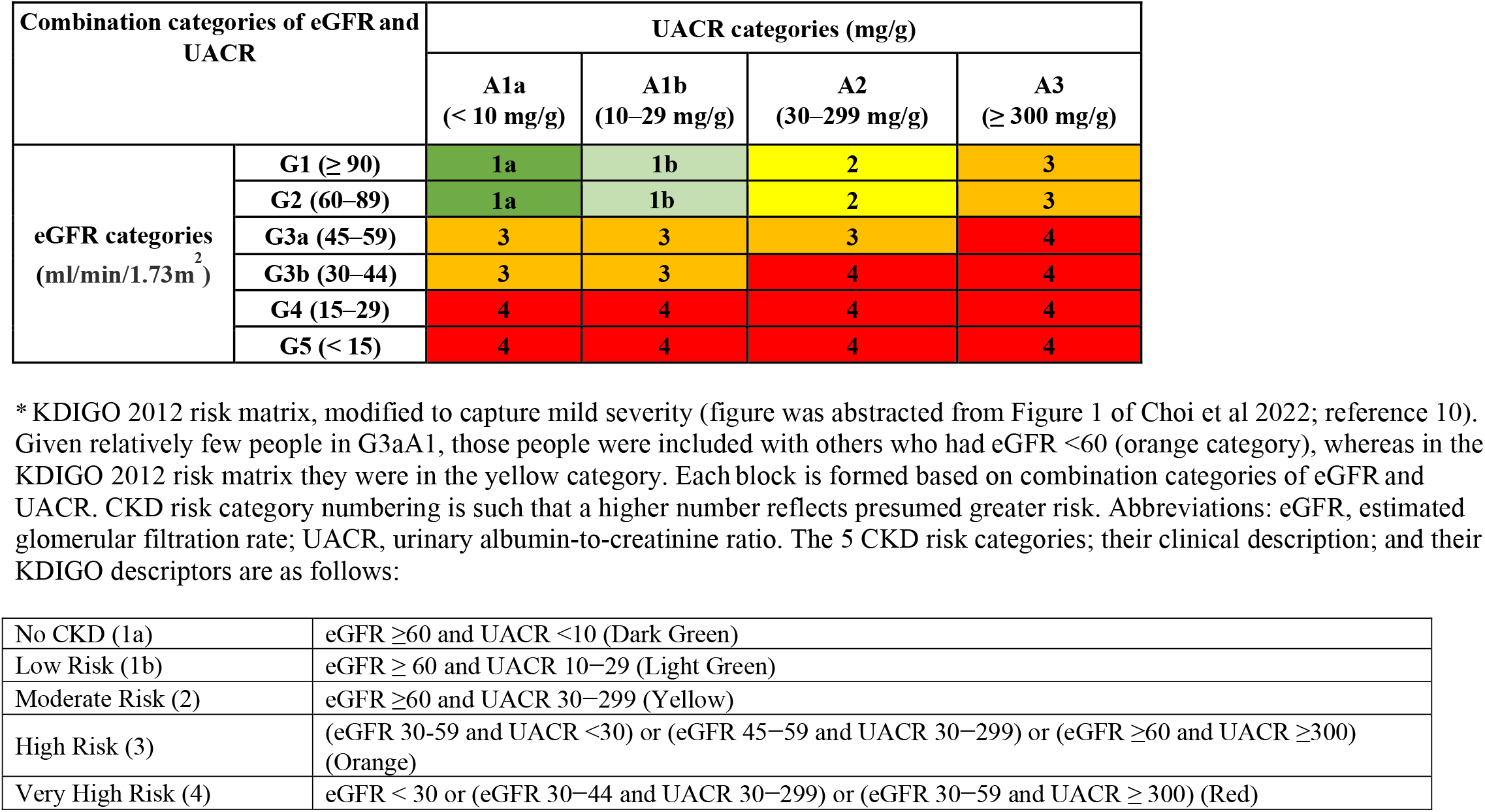
Definition of 5 chronic kidney disease (CKD) risk categories*.

